# Use of digital technology tools to examine patient adherence to a prescription omega-3 polyunsaturated acids therapy intended to mitigate cardiovascular risk: protocol and preliminary demographic data from the DIAPAsOn prospective observational study

**DOI:** 10.1101/2021.03.11.21252068

**Authors:** Gregory P. Arutyunov, Alexander G. Arutyunov, Fail T. Ageev, Tatiana V. Fofanova

## Abstract

Sustained adherence and persistence with prescription medications is considered essential to achieve maximal treatment benefit for patients with major chronic non-communicable diseases such as hyperlipidemia and lipid-associated cardiovascular disease. It is widely documented, however, that a large percentage of patients with these conditions have poor longer-term compliance with their treatments. The population of Russia is affected by poor compliance in the same ways as populations elsewhere and continues to have high rates of cardiovascular disease.

Hypertriglyceridemia is suspected of contributing to residual cardiovascular risk in patients whose low-density lipoprotein cholesterol (LDL-C) is well controlled. A preparation of highly purified omega-3 polyunsaturated fatty acids (EPA/DHA=1.2/1-90%, marketed as OMACOR), is indicated for the control of high triglyceride levels and also for secondary prevention of myocardial infarction (MI).

DIAPAsON is a prospective observational program designed to evaluate the impact of digital technologies on patient compliance and adherence in a large sample of patients whose treatment scheme includes EPA/DHA=1.2/1-90%. A bespoke electronic data capture system has been developed for this purpose that enables information obtained during clinic visits to be supplemented by remote patient self-reporting. Other objectives of the program include raising patients’ awareness about their condition via educational materials available in patient personal accounts in the electronic system.

This report outlines the rationale and methodology of DIAPAsOn and the primary demographics of the study population.

## Introduction

Low-density lipoprotein cholesterol (LDL-C), the primary lipoprotein component of total cholesterol (TC), is recognized as the most important lipid risk factor for coronary heart disease (CHD). An extensive array of clinical trials has demonstrated the value of lowering LDL-C as a means of prevention (especially secondary prevention) of major cardiovascular events, including myocardial infarction (MI) and stroke. These findings underpin the status of statins as first-line therapies to reduce cardiovascular risk via modulation of LDL-C levels. The Cholesterol Treatment Trialists (CTT) meta-analysis, which incorporated data from >90,000 patients in 14 randomized controlled trials, exemplifies the influence of LDL-C levels on cardiovascular risk: every 1 mmol/l (39 mg/dl) reduction in LDL-C leads to a decrease in overall mortality of 12%, a 19% reduction in coronary mortality and a 17% reduction in the incidence of stroke, with the effect size relating closely to the absolute reduction in LDL-C achieved and evident across the continuum of LDL-C levels [1,2].

Nevertheless, 5-year event rates in statin-treated patients in the CTT meta-analysis were 14% (compared with 18% in the reference group), and even higher residual risk was apparent in patients with pre-existing CHD or type 2 diabetes [1]. It has been estimated that some 50% of individuals who present with acute coronary syndrome (ACS) do not have elevated LDL-C, indicating that other factors account for a sizeable proportion of total cardiovascular risk [3].

The existence of residual risk in patients whose LDL-C is well controlled with medication [4,5] has focused attention on additional contributors to that persisting risk, including elevated plasma triglyceride (TG) levels [6]. Demonstration of a causal link between elevated TG and cardiovascular risk has been a matter of controversy, with evidence of an association between TG levels and cardiovascular risk often being attenuated after adjustment for other factors [7-11]. Considerations such as the reciprocal relation between levels of TG and levels of high-density lipoprotein cholesterol (HDL-C) (a correlation that persists even when TG are low) [12], as well as appreciation of qualitative metabolic interplay between TG-rich lipoproteins and HDL-C fractions, have nevertheless established a strong prima facie case for TG levels as one factor influencing residual cardiovascular risk in statin-treated patients, especially in patients characterized by a high TG:HDL-C ratio. Various national and international guidelines recognize that elevated TG may be implicated in the overall risk of CHD [13-15], notably so in patients with type 2 diabetes mellitus, obesity or metabolic syndrome [16-19].

Highly purified long-chain omega-3 polyunsaturated fatty acids (n-3 PUFAs), available as prescription-only medications, are approved in various countries for the management of elevated TG. These preparations are qualitatively distinct from dietary supplements of n-3 PUFAs [20]. One such prescription n-3 PUFA preparation (EPA/DHA=1.2/1-90%OMACOR, Market Authorization Holder Abbott Laboratories GmbH, Germany), is available in Russia as 1g capsules containing approximately 840 mg of eicosapentanoic acid and docosahexanoic acid and is approved for use at doses of 2–4 g/day for the regulation of TG. EPA/DHA=1.2/1-90%is also approved, at a daily dose of 1 g, for the secondary prevention of major cardiovascular events in patients who have survived an MI, an indication supported by the findings of a randomized clinical trial and corroborated by a recent Cochrane review [21].

Continuance with therapy is central to the attainment of full and sustained treatment benefit in cardiovascular prevention, as in a range of other major non-communicable diseases. Patients’ sense of ‘ownership’ of their situation, and the accompanying sense of empowerment, can be an important determinant of willingness to persevere with a course of therapy [22]. This can be a challenge when dealing with initially symptomless conditions such as hyperlipidemia, where the connection between symptomless aberrations in blood lipid levels and later major cardiovascular events can seem abstract.

The emergence of widely available digital and internet technologies with the potential to provide rapid or immediate bidirectional communication between healthcare professionals and patients may be an important new resource for promoting long-term adherence to therapies [23]. The DIAPAsOn study was designed to explore the impact of digital technology tools on patient adherence to therapy.

## Methods

### Overview

DIAPAsOn was a non-randomized, prospective, non-interventional observational study conducted at more than 100 centers in the Russian Federation to explore adherence to a prescription of EPA/DHA=1.2/1-90% as either a secondary preventive medical therapy for patients with a history of recent MI (approved dose 1g/day) or for blood lipid regulation in patients with endogenous hypertriglyceridemia that had proved insufficiently responsive to either dietary modification or other drug therapy (approved dose 2-4 g/day). As the objectives of the program are exploratory, there is no formal study hypothesis and no formal calculation of sample size: however, it was planned to enroll 3000 patients.

There is no control group in DIAPAsOn and patients do not undergo additional diagnostic tests or interventions other than those provided for by currently accepted standards of medical care.

A feature of DIAPAsOn is the use of digital technology to facilitate patient-initiated data collection. Patients have access to a customized electronic system that allowed them to enter data, including daily records of EPA/DHA=1.2/1-90% administration, and to complete various questionnaires relating to, for example, health-related quality of life (HRQoL) and product usability on a four-grade scale (very good, good, moderate, poor). If the reported grade is ‘moderate’ or ‘poor’, patients were asked to provide narrative details for this assessment.

This facility includes an option for patients to set up reminders to take the study medication. Patients are also able to report side effects in their personal accounts or to indicate if they had undergone any hospitalizations due to cardiovascular reasons, new cases of angina pectoris or non-fatal MI.

DIAPAsOn is registered at www.ClinicalTrials.gov (NCT03415152).

### Recruitment of study centers and patients

Selection of investigators and sites to participate in the program was based on the following requirements:

1. The ability to properly conduct the program, including capacity to complete an electronic case record form.
2. Existence within the study center of a cohort of adult (≥18 years) patients with a history of MI not earlier than 6 months previously for whom EPA/DHA=1.2/1-90% was prescribed as part of a medical secondary prevention regimen and/or existence within the study center of a cohort of patients with a diagnosis of hypertriglyceridemia not adequately controlled by a hypolipidemic diet.

Patients provisionally satisfying the criteria set out in (2) were eligible to participate in DIAPAsOn if they had been taking EPA/DHA=1.2/1-90%for no more than 14 days at the time of enrolment and if they were considered capable, either personally or with the assistance of immediate relatives, of submitting data through a mobile phone app or web browser.

Candidates for enrolment were excluded if they were taking medications other prescription-only omega-3 PUFA products or nutrition supplements containing omega-3 PUFAs at screening or had taken such compounds within the previous 6 months. Other exclusion criteria comprised:

- Pregnancy or breastfeeding.
- Known sensitivity to the active substance, excipients or soy.
- Exogenous hypertriglyceridemia (Frederikson type I hyperchylomicronemia).
- Current participation in any other clinical or observational study, or involvement within the previous 30 days.
- Any other clinical state that, in the opinion of the center investigator, made the patient unsuited to inclusion.

### Objectives and endpoints

The primary objective of DIAPAsOn was to assess adherence to therapy with EPA/DHA=1.2/1-90% in post-MI patients or patients with hypertriglyceridemia.

Secondary goals included evaluation of:

- reasons for termination of the study therapy
- in-study HRQoL
- health outcomes in the different patient subgroups.

### Schedule of visits and data collection

Three principal clinic visits featured in the timeline of the DIAPAsOn study, as illustrated in **Figure 1**. At each of these, patients were interrogated regarding their adherence to EPA/DHA=1.2/1-90% and their compliance with that therapy was represented by answers to the Questionnaire of Treatment Compliance [24]. This instrument, which has been used to investigate compliance with antihypertensive medication among Russian patients, produces a numerical indication of compliance, as follows: 12–15 points, very high; 8–11 points, high; 4–7, moderate; 0–3, low.

**Figure 1.**
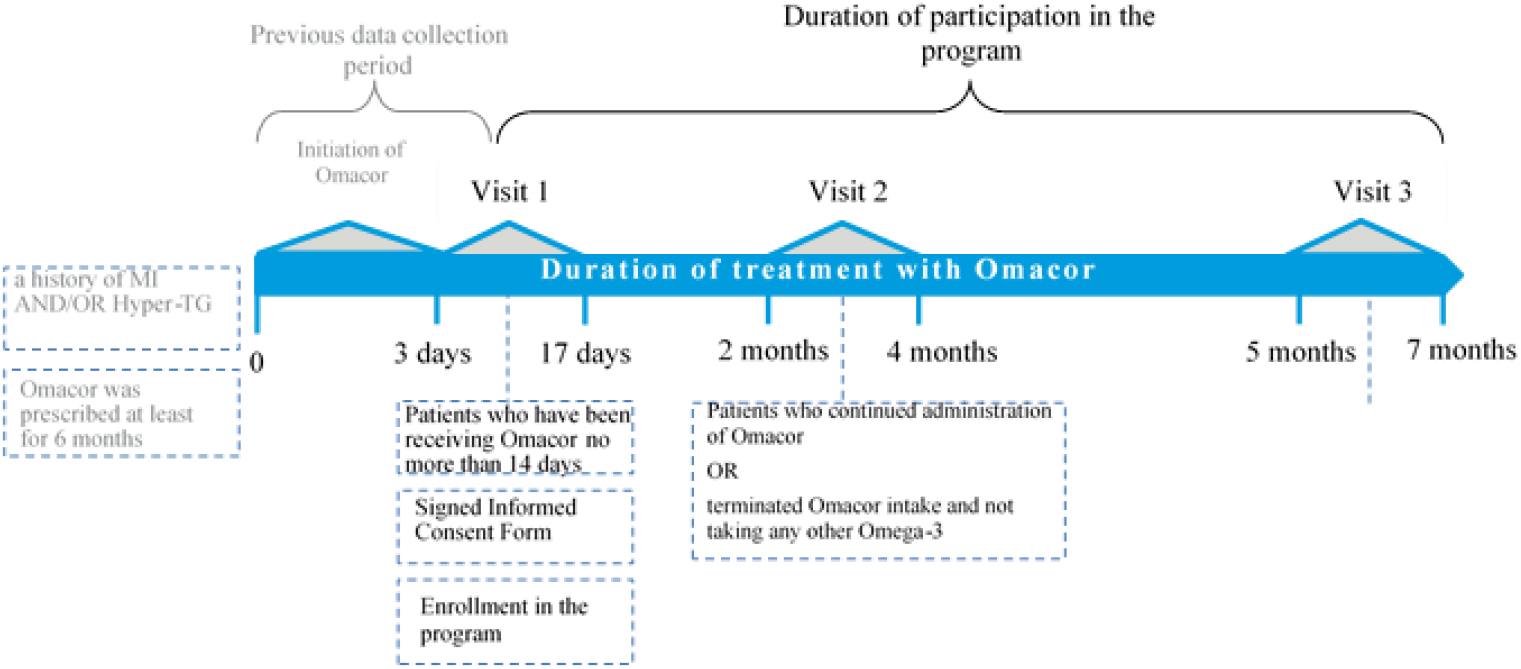
Design and chronology of the DIAPAsOn study.

Other information elicited or obtained at these visits included: significant concomitant therapy, blood pressure and heart rate. Because EPA/DHA=1.2/1-90% was administered within the frames of normal medical practice, rather than being supplied by the sponsor, information about the batch number and shelf life of EPA/DHA=1.2/1-90% were also recorded at each visit.

Blood lipid profile was determined at each clinical presentation. As DIAPAsOn was a non-interventional observational study, no central laboratory procedures were performed. All laboratory tests were conducted in accordance with routine local clinical practice.

Safety and adverse events data were elicited at each visit. In the event of a patient discontinuing the study, enquiries were made about the reasons for this, under the following headings: adverse drug reactions; lack of effect; inconvenience of use; non-availability in pharmacies; other.

The principal clinic visits were supplemented by inter-visit phone calls focused on adherence to therapy and safety.

### Mobile patient engagement technology

The electronic patient engagement and data collection system used in DIAPAsOn was developed in collaboration with the medical online platform ROSMED.INFO (Moscow, Russian Federation; www.rosmed.info), which has extensive experience in the development and operation of such facilities in the Russian Federation, including: the Russian Severe Asthma Registry (RSAR; NCT03608566); the all-Russian register of patients with hearing impairment and the integrated support program (anticipated recruitment 12 million people); and the Russian Register of patients aged >80 years with ACS. ROSMED.INFO is also involved in the maintenance, on behalf of the Russian Glaucoma Society, of the first Russian pharmaco-epidemiological study of the treatment of glaucoma patients with retinoprotective interventions and adherence to therapy.

The system devised for DIAPAsOn is configured to minimize technical and ergonomic barriers to participant adoption and use. Contributing patients may work with the web version (without installing software on a local PC) or may download the relevant app for IOS (Apple) or Android smartphones; all versions of the online platform are configured to work with all popular browsers.

The ROSMED.INFO platform as used in DIAPAsOn is a software product that satisfies current requirements of Russian legislation pertaining to the management of personal data and the implementation of observational studies, primarily Russian Federal Law No. 152.

The ROSMED.INFO platform is included in the Russian Unified Register of computer programs and databases of the Ministry of Communications of Russia. Certification of quality management and information security management is conducted in accordance with ISO 9001: 2015 and ISO/IEC 27001: 2013.

Operational domains of the digital platform are depicted in **Figure 2** and differentiate between an “administrative tier” occupied by the service manager and the study sponsor and an “executive” tier that include dedicated areas (“cabinets”) for use by patients and physicians.

**Figure 2.**
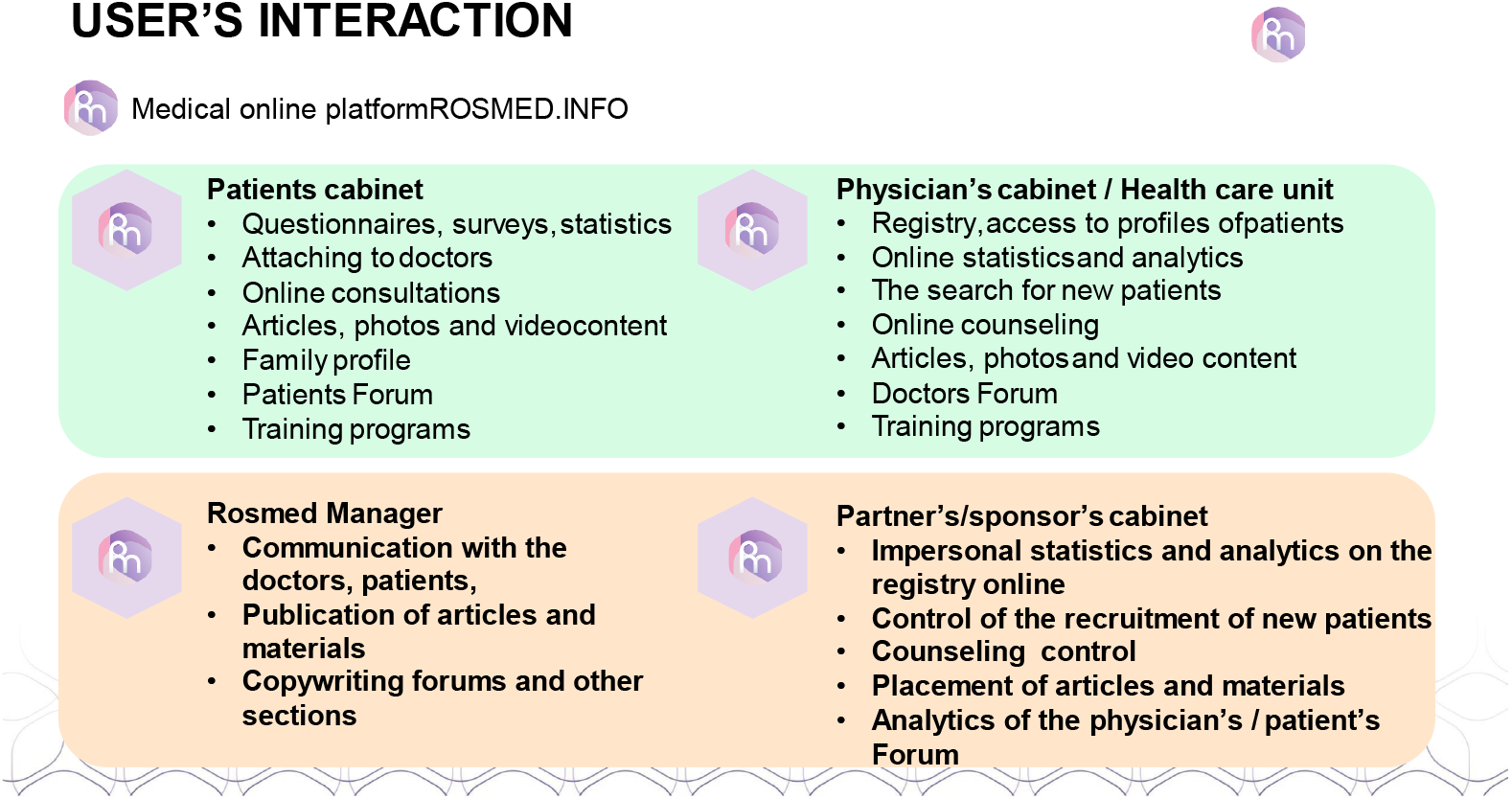

Specimens of the Physician Cabinet GUI are shown in **Figures 3 & 4** along with a bulleted summary of the facilities available to physicians **(Figure 3)**. (These screengrabs have been translated into English for the convenience of an international readership: the working versions are in Russian and are available on application to the author for correspondence. The examples shown here have also been anonymized.)

**Figure 3.**
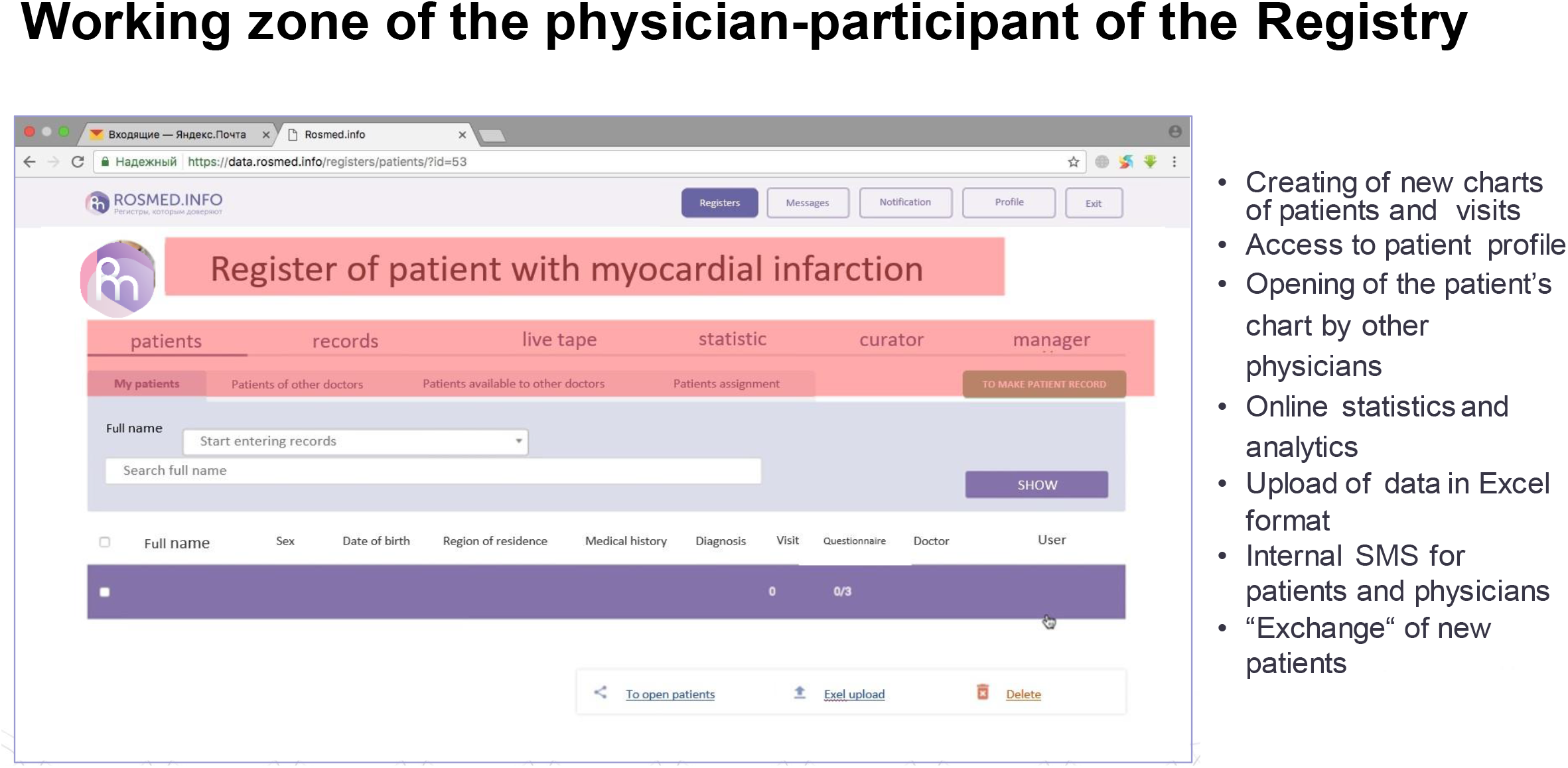

**Figure 4.**
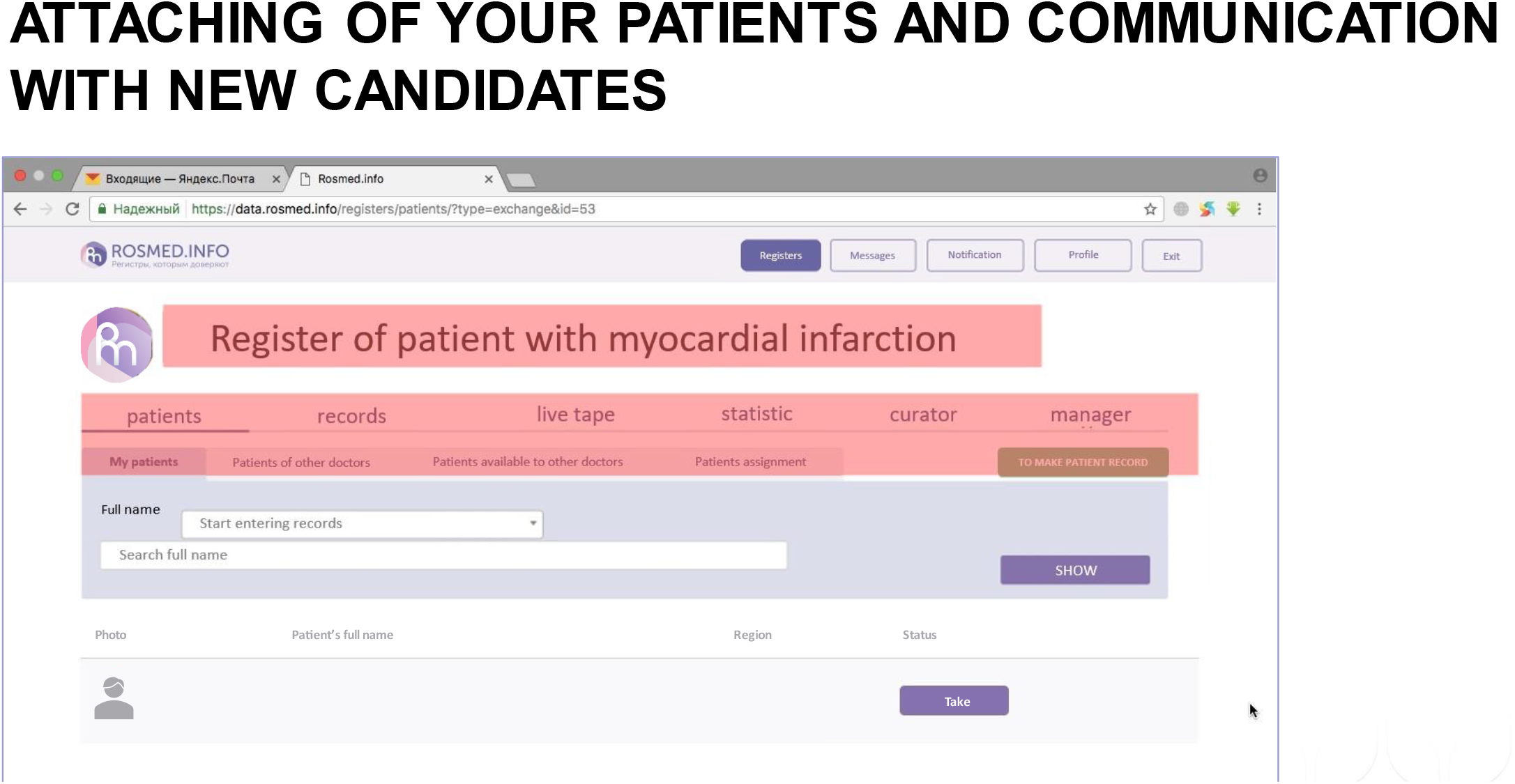

The landing page for the Patient Cabinet is illustrated in **Figure 5** and identifies 5 main drop-down menus designed to allow patients easy access to the full range of facilities and services provided by the technology. These include access to notices and push-reminder services **(Figure 6)** and an internal social network **(Figure 7)**. The patient registration page has deliberately been kept simple to minimize set-up friction and encourage patient enrolment **(Figure 8)**. (These screengrabs have been translated into English for the convenience of an international readership: the working versions are in Russian and are available on application to the author for correspondence. The examples shown here have also been anonymized.)

**Figure 5.**
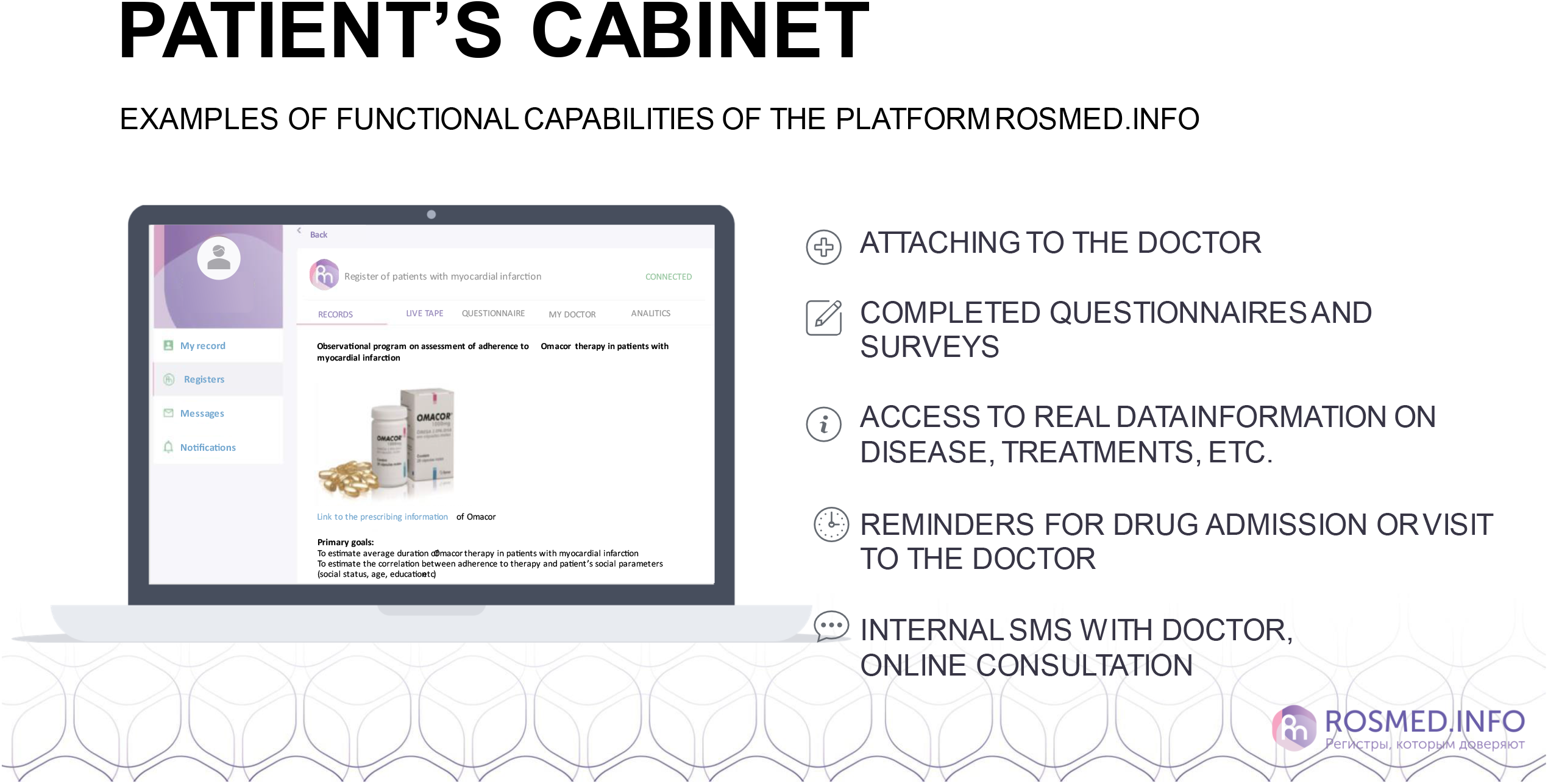

**Figure 6.**
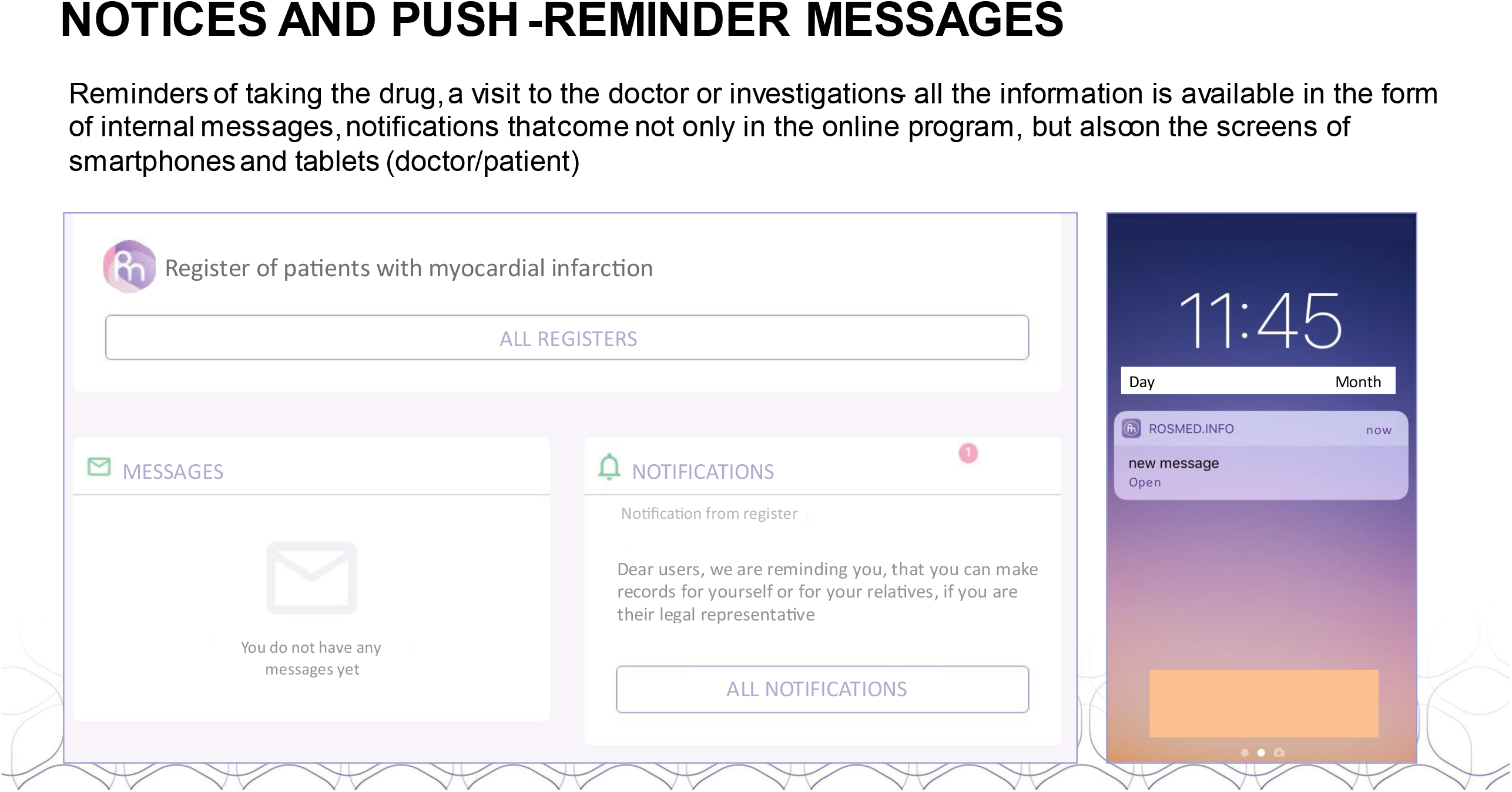

**Figure 7.**
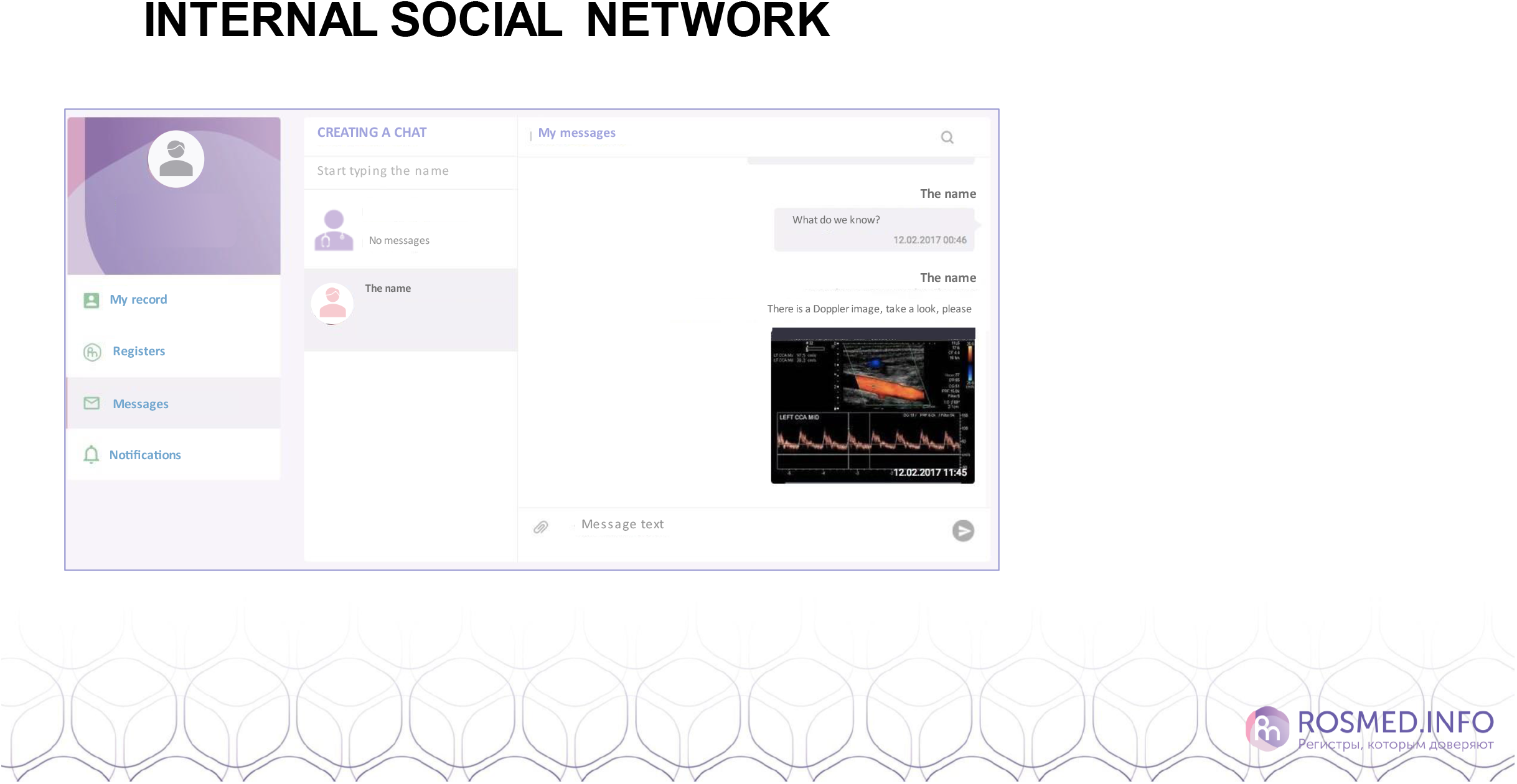

**Figure 8.**
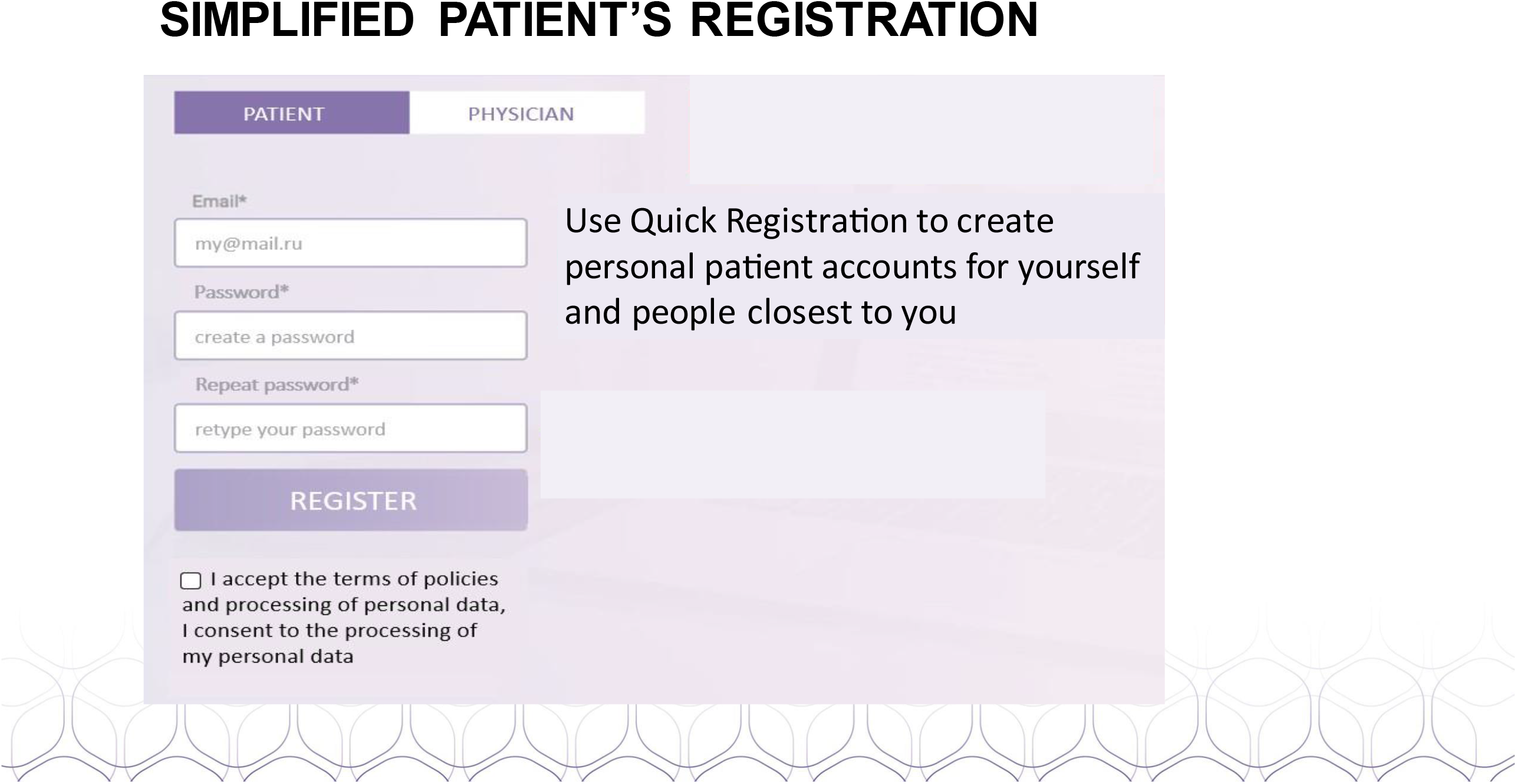

### Statistical and analytical plans

Statistical analysis is performed in accordance with a pre-approved Statistical Analysis Plan, using the statistical programming language R (version 3.4.3. https://mran.microsoft.com/releases/3.4.3).

As DIAPAsOn is an open, non-randomized observational program, statistical methods are planned to be mainly descriptive.

Comparison of patient data between scheduled clinical visits is carried out using tudent’s t-test for dependent samples, or c emar’s test for qualitative data or all comparisons, statistical significance is calculated. For applicable endpoints, further analysis is performed in subgroups of patients with different adherence rates at Visits 2 and 3.

The primary endpoint is assessed in the per-protocol population, which includes all patients for whom data are collected, at least at Visit 2. The safety population includes all patients who complete at least Visit 1. This population was used to record reports of adverse drug reactions (ADR), serious ADRs (SADR), and other safety information.

Analysis of the primary endpoint includes (1) determination of the mean adherence rate, calculated as the sum of days when the patient took the full prescribed dose of EPA/DHA=1.2/1-90% during the specified period divided by the total number of days in that period, at the end of the study (Visit 3); and (2) mean score on the National Questionnaire of Treatment Compliance, determined at the end of the study (Visit 3).

### Ethics

Ethical oversight of the DIAPAsOn study is exercised by the independent Interuniversity Ethics Committee (IEC; Moscow, Russian Federation). Initial written approval of the IEC was issued on 19 October 2017, before the start of the study. The study was then implemented in accordance with the protocol version of 8 November 2017.

DIAPAsOn conforms to the requirements of Good Clinical Practice (GCP) and to all applicable national standards relevant to the rights, safety and well-being of all study participants in accordance with the provisions of the Declaration of Helsinki and all relevant national legislation and related provisions. Advanced informed written consent is obtained from the patient to use and/or disclose personal and/or medical information. Prospective patients are apprised of their right to decline further participation in DIAPAsOn at any time and for any declared reason (or for no reason) without prejudice to any subsequent treatment.

Individual center investigators are responsible for ensuring quality control, execution of the program and the collection, documentation and submission of data in accordance with the protocol, standards of GCP and all applicable local laws.

### Administrative structure

The tripartite functional organization of the project is illustrated in **Figure 9**. The study principal investigator is Professor Grigory Pavlovich Arutyunov, Pirogov Russian National Research Medical University, Moscow, Russian Federation. A list of investigators at each of the 108 participating centers is presented in **Supplementary Table 1**.

**Figure 9.**
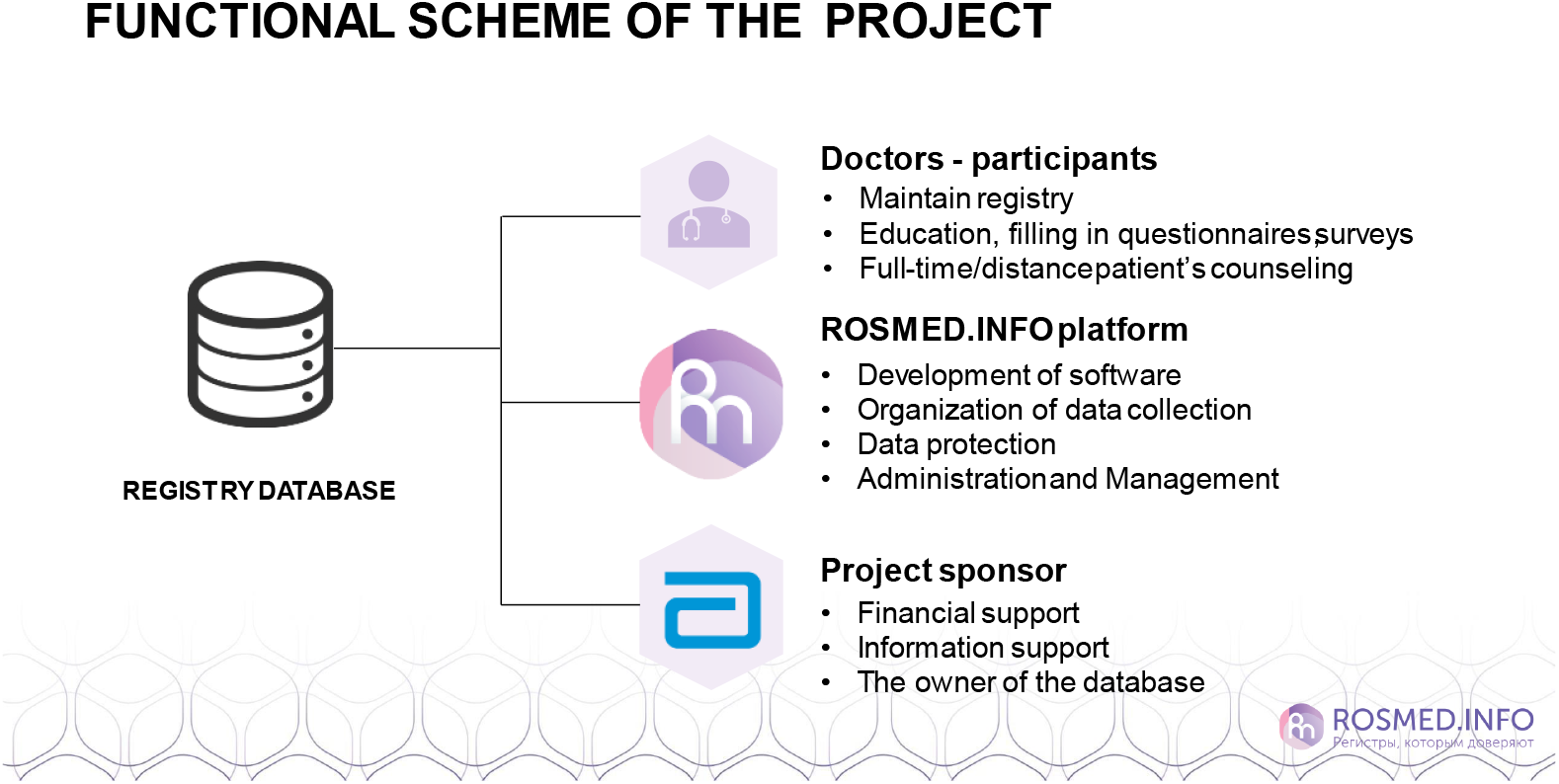

ROSMED.INFO is responsible for the development and maintenance of software for the digital data collection and adherence facilities, and for data collection and protection.

Contract research services are provided by RSMI LLC (Moscow, Russian Federation), and data statistical services by Sciencefiles LLC (Yekaterinburg, Russian Federation).

DIAPAsOn is sponsored by Abbott Laboratories (Moscow, Russian Federation), which is the legal owner of the registry database of digital information generated during the study.

## Results

A total of 3000 patients were initially included in the program but 428 (14.3%) were excluded from the program because Visit 1 data were incomplete. Valid and complete data from Visit 1 were available for 2572 patients (85.7%) who constituted the safety population, and data from a per-protocol contingent of 2167 patients was accrued, 405 patients having been lost due to early termination (i.e. before Visit 3).

Within the overall per-protocol population, 898 patients (41.4%) were prescribed EPA/DHA=1.2/1-90% for secondary prevention after MI and 1269 (58.6%) were prescribed EPA/DHA=1.2/1-90% for hypertriglyceridemia. The program was completed per protocol by 1975 patients, 780 of whom were being treated for secondary prevention and 1195 of whom were being treated for hypertriglyceridemia.

Because DIAPAsOn is an observational study, the protocol stipulated that discontinuation of EPA/DHA=1.2/1-90% is not a reason to withdraw a patient from the study.

Detailed tabulations of the demographic profile of the DIAPAsOn patients are provided in **Tables 1 & 2**, sub-stratified according to the reason for treating with EPA/DHA=1.2/1-90% (i.e. secondary prevention post-MI or management of hypertriglyceridemia). In summary, men account for 52.84% of the total population and 67.7% of the subgroup prescribed EPA/DHA=1.2/1-90% for secondary prevention post-MI (n=608); conversely, most of the patients prescribed EPA/DHA=1.2/1-90% for hypertriglyceridemia are women (n=732; 57.68%). The study population is almost exclusively Caucasian (97.74%) and almost all other patients (n=43) were classified as Asian.

**Table 1.** Age, height, weight and BMI data for the overall population and designated subgroups (based safety population with data available from Visit 1; N = 2572).

| Parameter | Age (years) | Height (m) | Weight (kg) | BMI (kg/m<sup>2</sup>) |
| --- | --- | --- | --- | --- |
| Mean | 59.86 | 1.71 | 87.09 | 29.89 |
| SD | 11.62 | 0.09 | 16.85 | 5.62 |
| Min | 20 | 1.45 | 42 | 18.42 |
| Max | 93 | 2.01 | 195 | 62.16 |

Secondary prevention subgroup (N = 1171)
| Parameter | Age (years) | Height (m) | Weight (kg) | BMI (kg/m<sup>2</sup>) |
| --- | --- | --- | --- | --- |
| Mean | 61.79 | 1.73 | 85.96 | 28.54 |
| SD | 10.29 | 0.09 | 15.14 | 4.08 |
| Min | 27 | 1.48 | 42 | 18.42 |
| Max | 93 | 2 | 165 | 50.36 |

Hypertriglyceridemia subgroup (N = 1401)
| Parameter | Age (years) | Height (m) | Weight (kg) | BMI (kg/m<sup>2</sup>) |
| --- | --- | --- | --- | --- |
| Mean | 58.24 | 1.69 | 88.04 | 31.01 |
| SD | 12.41 | 0.08 | 18.11 | 6.43 |
| Min | 20 | 1.45 | 49 | 18.67 |
| Max | 89 | 2.01 | 195 | 62.16 |

**Table 2.** Sex, education, employment, marital and smoker status for the per-protocol population (N = 2167), subdivided by reason for prescribing EPA/DHA=1.2/1-90%.

|  | Overall population |  | Secondary prevention post-MI |  | Hypertriglyceridemia |  |
| --- | --- | --- | --- | --- | --- | --- |
|  | N | Proportion (%) | N | Proportion (%) | N | Proportion (%) |
| **Sex** |  |  |  |  |  |  |
| Male | 1145 | 52.84 | 608 | 67.71 | 537 | 42.32 |
| Female | 1022 | 47.16 | 290 | 32.29 | 732 | 57.68 |
| **Education** |  |  |  |  |  |  |
| Secondary education drop-out | 17 | 0.78 | 13 | 1.45 | 4 | 0.32 |
| Secondary general | 238 | 10.98 | 86 | 9.58 | 152 | 11.98 |
| Secondary vocational | 673 | 31.06 | 280 | 31.18 | 393 | 30.97 |
| Higher | 1199 | 55.33 | 504 | 56.12 | 695 | 54.77 |
| Supplementary vocational | 40 | 1.85 | 15 | 1.67 | 25 | 1.97 |
| **Work status** |  |  |  |  |  |  |
| Working | 1199 | 55.33 | 445 | 49.55 | 754 | 59.42 |
| Not working | 968 | 44.67 | 453 | 50.45 | 515 | 40.58 |
| **Marital status** |  |  |  |  |  |  |
| Single | 91 | 4.2 | 33 | 3.67 | 58 | 4.57 |
| Married | 1744 | 80.48 | 669 | 74.5 | 1075 | 84.71 |
| Divorced | 124 | 5.72 | 60 | 6.68 | 64 | 5.04 |
| Widowed | 208 | 9.6 | 136 | 15.14 | 72 | 5.67 |
| **Smoker status** |  |  |  |  |  |  |
| Smoker | 613 | 28.29 | 284 | 31.63 | 329 | 25.93 |
| Non-Smoker | 1554 | 71.71 | 614 | 68.37 | 940 | 74.07 |

Patients’ mean age is 59.86±11.62 years, and overall mean body mass index (BMI) is 29.9±5.6 kg/m^2^. Patients in the secondary prevention subgroup are slightly older than those enrolled for hypertriglyceridemia (61.79±10.29 vs. 58.2±12.4 years), whereas the latter have a higher average BMI (31.0±6.4 vs. 28.5±4.1 kg/m^2^). Just over a quarter of all patients (28.29%) are recorded as active smokers, including 31.63% of patients in the secondary prevention subgroup (hypertriglyceridemia subgroup, 25.9%). In the secondary prevention subgroup the average time since the index MI to start of study therapy with EPA/DHA=1.2/1-90% is 3.04 ± 1.73 months. The most frequently recorded concomitant diseases identified in the study population at baseline (based on International Classification of Diseases, version 10 codes) comprised ‘ ypertensive diseases’ (I10–I15), recorded in 63.82% of patients. Within those categories, ‘ ypertensive heart disease without congestive heart failure’ (33 5% of patients, n=1383) and ‘Essential hypertension’ (15 1% of patients, n=328) are conspicuous ‘ oronary heart disease’ (I20–I25) is recorded in 17.9% of patients (n=388), ‘ ther heart disease ’ (I30-I52), including unspecified atrial fibrillation and atrial flutter, in 16 22 % (n=352) and ‘ iabetes mellitus’ in 7 11% of patients (n=1564) ‘ etabolic disorders’ (E70 – E90) are recorded in 10.8% of patients (n=235).

Therapies prescribed during the 12 months preceding enrolment are dominated numerically by lipid-modifying agents, taken by 1600 patients (73.83%). The single most frequently prescribed drug in this category is rosuvastatin (n=906, 41.8 % of the per-protocol population), followed by atorvastatin (n=601, 27.7 % %). Combinations of lipid-modifying drugs accounted for a further 6.9% (n=150). Just over half the patients (n=1140, 52.6 %) have been prescribed anti-platelet agents, primarily aspirin (n=583), and 40.6 % (n=879) have been prescribed beta-blockers, predominantly bisoprolol (n=586). A further 694 patients (32%) received angiotensin-converting enzyme (ACE) inhibitors, either as monotherapy (n=438) or in combinations with other classes of antihypertensive drugs, including diuretics and calcium-channel blockers. Perindopril is the most widely prescribed ACE inhibitor (n=325).

Variations are apparent in the two indication-specific subgroups. Thus, among patients who received EPA/DHA=1.2/1-90% for secondary prevention post-MI, use of anticoagulants is recorded in 96.4%, use of beta-blockers in 62.9%, use of ACE inhibitors in 31.9% and use of lipid-modifying agents in 73.6%. Pre-study use of lipid-modifying agents is at a similar level in the subset of patients with hypertriglyceridemia (74.0%) but use of the other classes of agents specified above was at a lower level. Baseline mean blood pressure is 133.7±17.3/81.4±8.8 mmHg. In the cohort overall, and in the hypertriglyceridemia subset, some 25% of patients are considered to have clinically relevant elevation of systolic blood pressure, compared with 18% of those in the post-MI subgroup. Approximately 13% in both subgroups are considered to have clinically relevant elevation of diastolic blood pressure.

Full analysis of the findings of DIAPAsOn in respect of the influence of patient-facing digital engagement and reporting technologies on adherence to EPA/DHA=1.2/1-90% therapy is in progress and is expected to be the subject of a separate report.

## Discussion

Rates of death from CHD have been falling in Russia in recent decades but the rate of decline has been less marked than in other European countries and age-standardized mortality rates remain markedly higher than in other nations [25, 26]. Data from the Know Your Heart survey and other investigations suggest that not all of this discrepancy is explained by blood lipids (alcohol appears to exert a notable influence) but lipids are regarded as central determinants of risk [27,28]. Additional efforts to improve the management of coronary risk are therefore warranted and a focus on patient adherence to secure maximum benefit from available therapies is one logical dimension of that response.

It is clear from the results of the CEPHEUS II study that failure to reach targets for lipid-based risk reduction is widespread in Russia [29]. Patient-related factors associated with non-attainment of targets identified in that study included considering it acceptable to miss prescribed doses more than once per week.

Two-thirds of the patients who participated in DIAPAsOn have concomitant hypertension. Recently published results from the ANICHKOV trial of patients in Russia with both high blood pressure and high cholesterol illustrate the necessity of non-statin therapy in patients with high TG and the difficulties of attaining target levels for both lipids and blood pressure [30].

In response to these deficits, we have developed digital technology tools designed to stimulate patient engagement and education and have recruited 3000 patients to a first trial of the effectiveness of this mobile health technology. We believe this research in preventive cardiovascular medicine to be the first of its kind undertaken at scale in Russia: it may provide a model for further developments in this field.

Limitations of DIAPAsOn must be recognized. As with observational studies in general, the absence of a control group precludes any determination of cause and effect and the potential for biases in any trial of this type must be acknowledged. A retrospective calculation of the Nichol score for DIAPAsOn confirmed that our study rated favorably in the subcategories of ‘ isease-elated riteria’ and ‘ ompliance Definition and Measurement Criteria’ but scored less strongly in ‘ tudy esign riteria’ he duration of follow-up is appropriate for a pilot trial of a technical innovation but a substantially longer period of observation will be needed to demonstrate if our strategy for digital patient engagement translates into a robust, sustained and meaningful improvement in long-term compliance and adherence. The online resources are all developed and presented in Russian and the study population is ethnically highly homogeneous: those factors may restrict the generalizability of findings from this study. We have not performed a pre-study evaluation of the online services used in DIAPAsON using, for example, the Mobile App Rating Scale (MARS) and there is no pilot testing phase.

## Data Availability

Not applicable to this submission.

## Conclusions

DIAPAsOn is a prospective study of a digital technology and patient self-reporting platform to enhance adherence with EPA/DHA=1.2/1-90% therapy in patients who have experienced a first MI or who have a diagnosis of hypertriglyceridemia. Experience and insight from this study may inform the further development of mobile health engagement technologies for Russian populations to improve compliance and adherence with medications and thereby reduce the risk of clinical cardiovascular events.

## Acknowledgements

The investigators thank the Ethics Committee of Pirogov Russian National Research Medical University Ethical for its advice and guidance on the development of the research protocol.

Manuscript preparation was assisted by **Hughes associates**, Oxford, UK.

---

**Supplementary Table 1.**
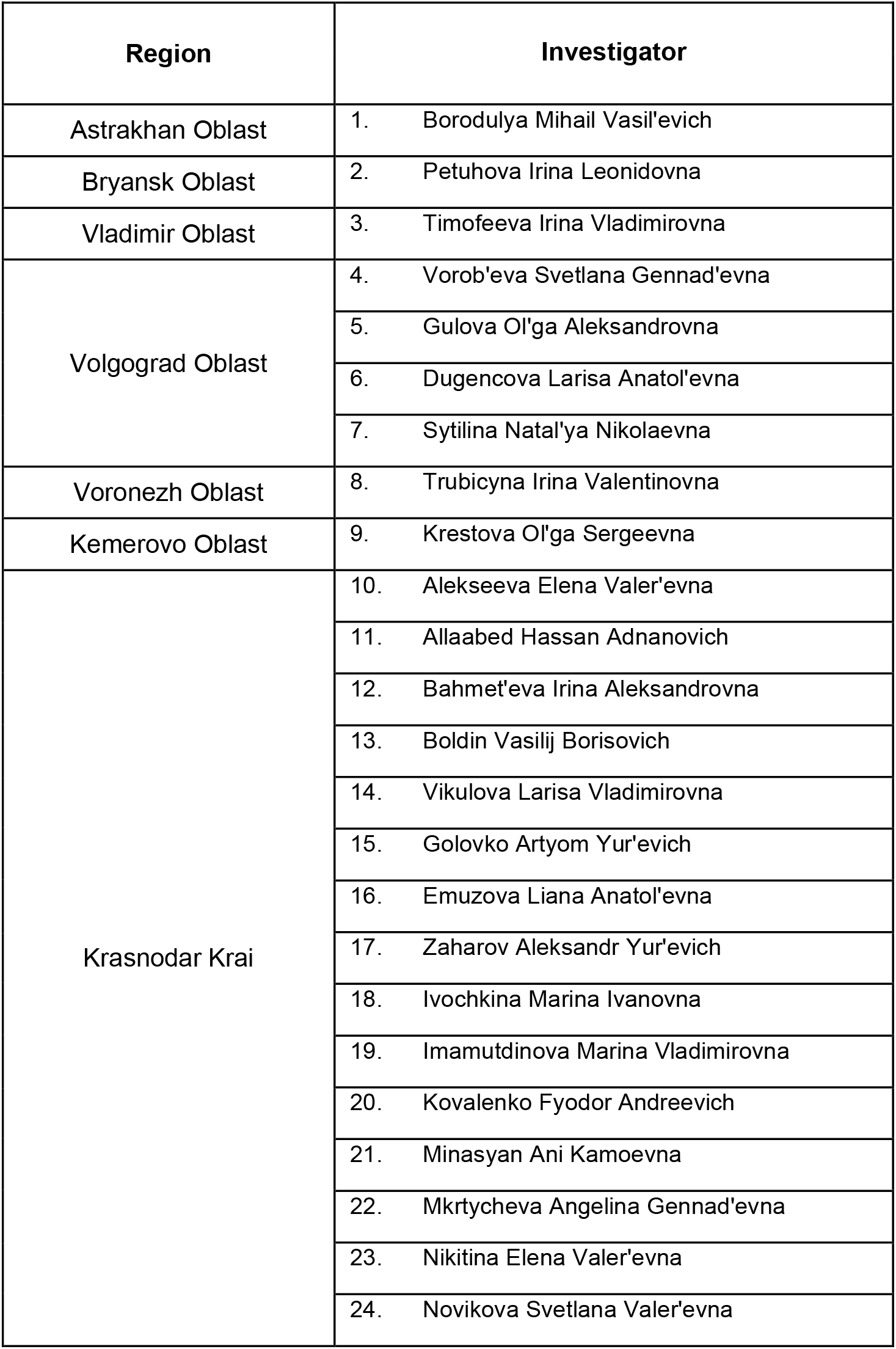
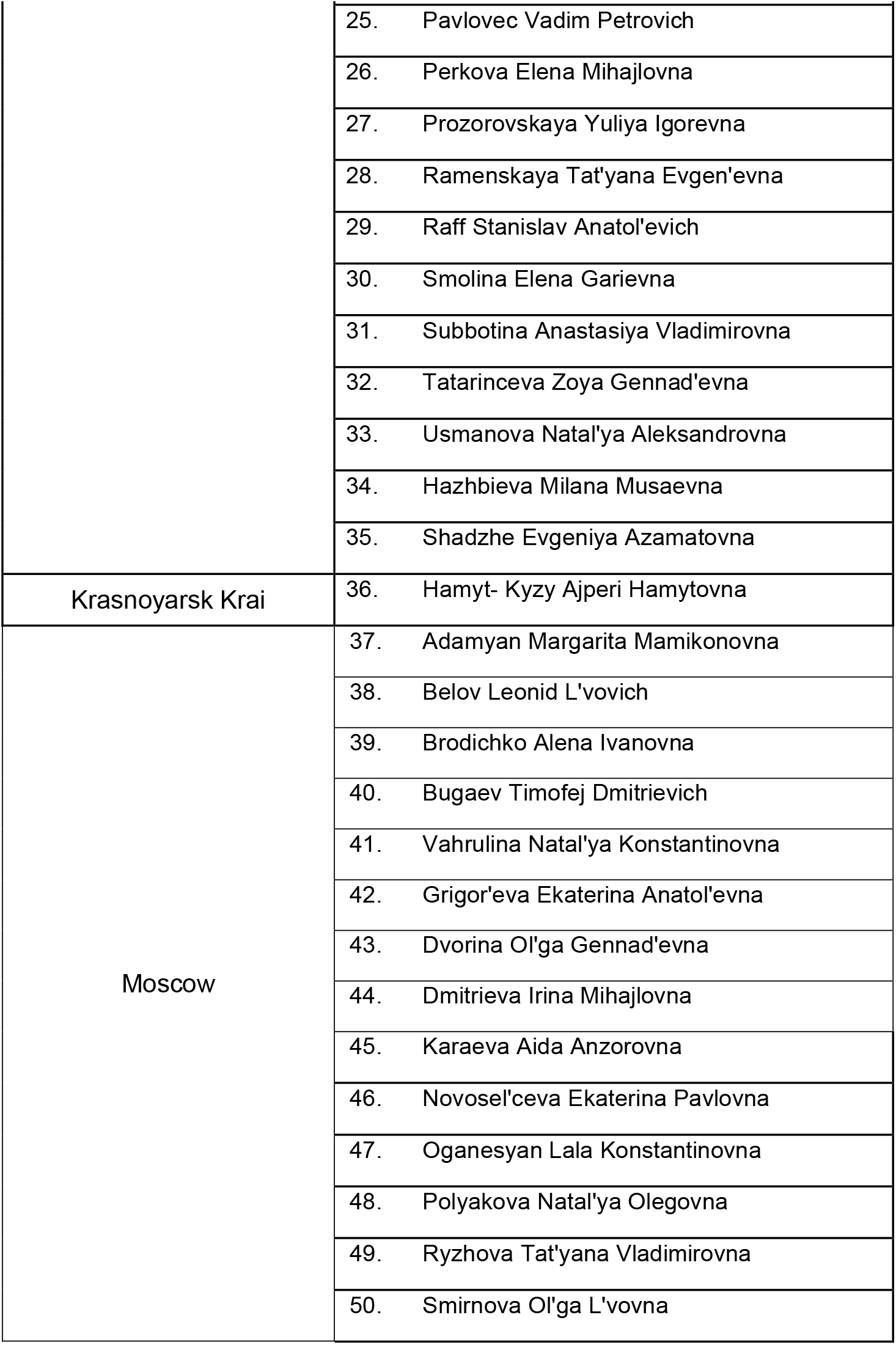
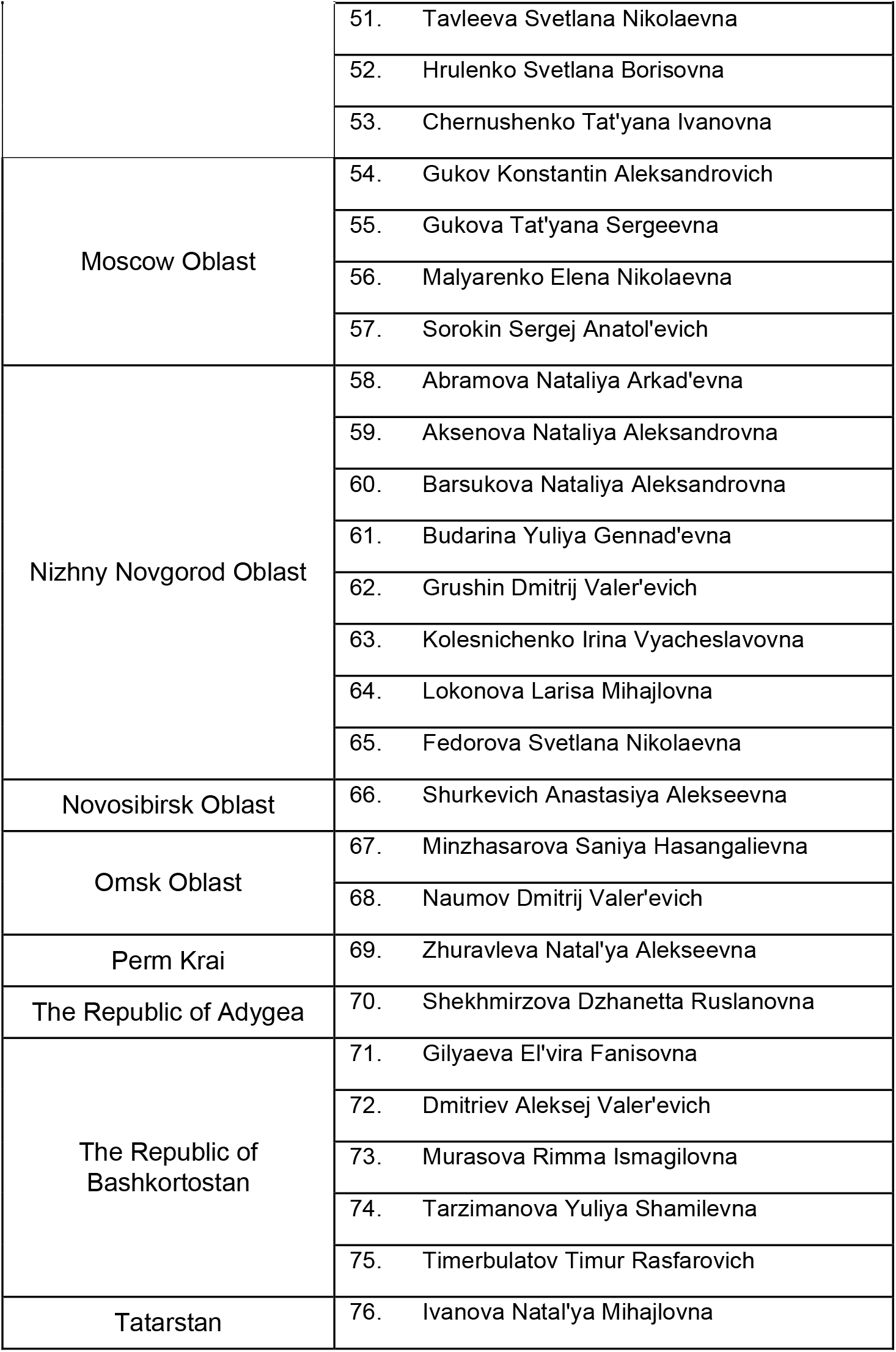
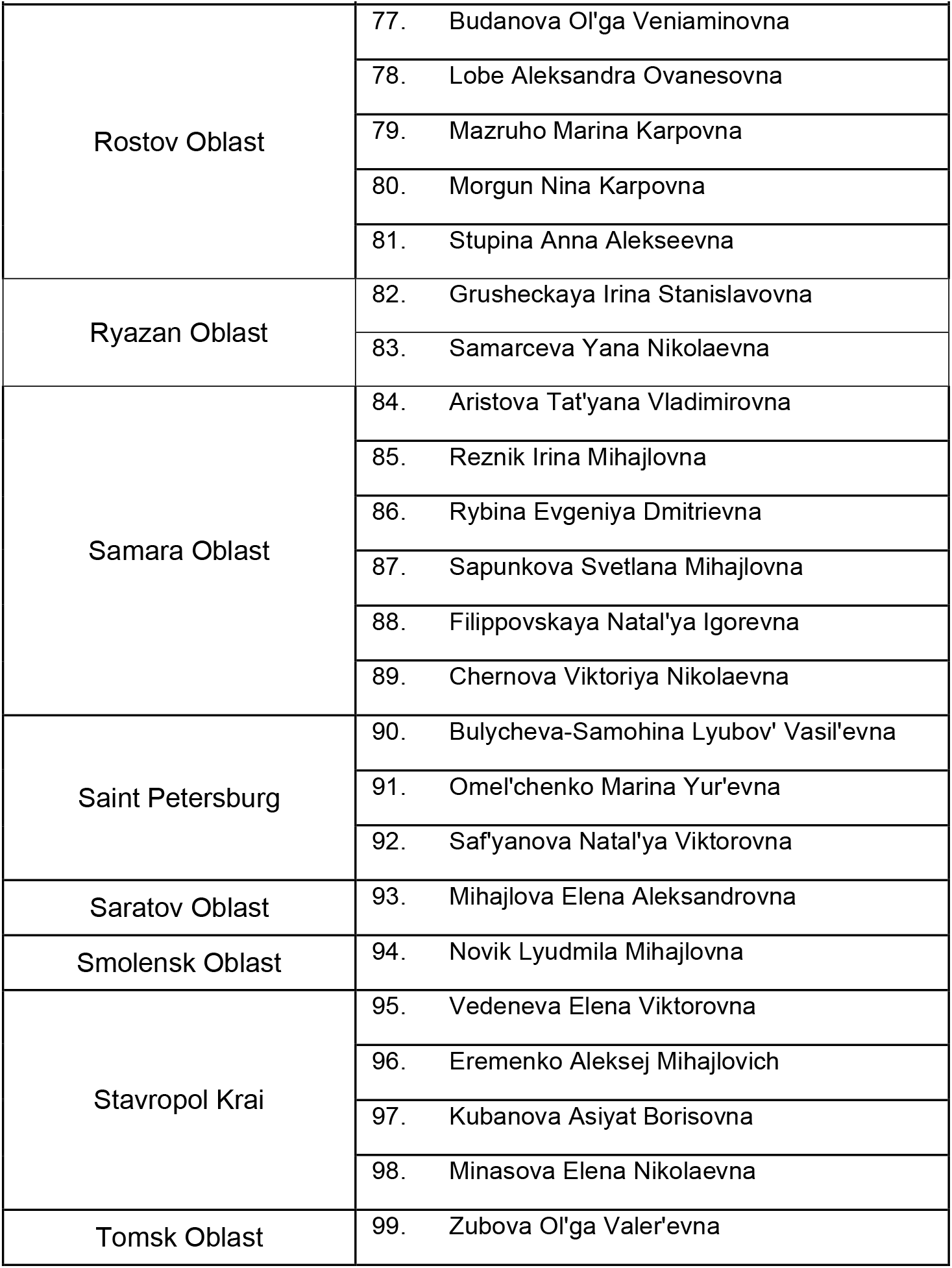
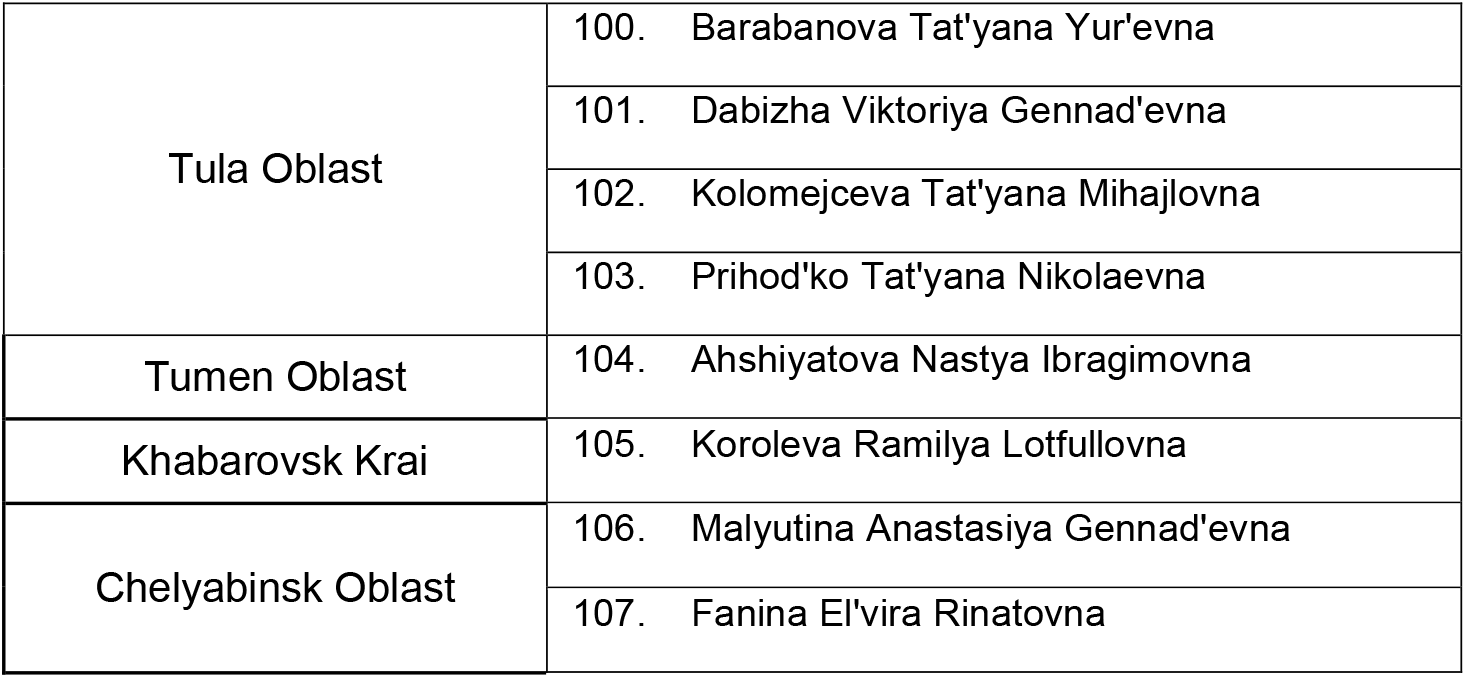
Acknowledgement of DIAPASoN centre investigators.

